# Missing data and missed infections: Investigating racial and ethnic disparities in SARS-CoV-2 testing and infection rates in Holyoke, Massachusetts

**DOI:** 10.1101/2023.05.24.23290470

**Authors:** Sara M. Sauer, Isabel R. Fulcher, Wilfredo R. Matias, Ryan Paxton, Ahmed Elnaiem, Sean Gonsalves, Jack Zhu, Yodeline Guillaume, Molly Franke, Louise C. Ivers

## Abstract

Routinely collected testing data has been a vital resource for public health response during the COVID-19 pandemic and has revealed the extent to which Black and Hispanic persons have borne a disproportionate burden of SARS-CoV-2 infections and hospitalizations in the United States. However, missing race and ethnicity data and missed infections due to testing disparities limit the interpretation of testing data and obscure the true toll of the pandemic. We investigated potential bias arising from these two types of missing data through a case study in Holyoke, Massachusetts during the pre-vaccination phase of the pandemic. First, we estimated SARS-CoV-2 testing and case rates by race/ethnicity, imputing missing data using a joint modelling approach. We then investigated disparities in SARS-CoV-2 reported case rates and missed infections by comparing case rate estimates to estimates derived from a COVID-19 seroprevalence survey. Compared to the non-Hispanic white population, we found that the Hispanic population had similar testing rates (476 vs. 480 tested per 1,000) but twice the case rate (8.1% vs. 3.7%). We found evidence of inequitable testing, with a higher rate of missed infections in the Hispanic population compared to the non-Hispanic white population (77 vs. 58 infections missed per 1,000).

## BACKGROUND

Research has demonstrated the disproportionate impact of the COVID-19 pandemic on Black, Indigenous, and Hispanic populations in the United States [1-3]. These communities experienced higher rates of SARS-CoV-2 infection, hospitalization, and COVID-19-related mortality compared to non-Hispanic white populations [4-11] – a result of structural racism, whereby systems, policies, and practices have created racial inequities in employment, housing, healthcare, and wealth [12]. Routinely collected COVID-19 testing data has been a vital resource to investigate racial and ethnic disparities in COVID-19-related outcomes. However, a full understanding has been limited by incomplete race and ethnicity data [13] and the absence of data for infected, but untested, individuals [3].

Missing data on race and ethnicity is common in US COVID-19 testing databases [13,14]. As of August 1, 2020, US laboratories were required to report race and ethnicity for all COVID-19 tests [15]. Despite this, 32% of reported cases had missing information on race and 42% on ethnicity between August 1 and December 31, 2020 [16]. Recent studies investigating racial and ethnic disparities typically exclude participants with missing information or group them into an “unknown” category for analysis [3,17]. This can underestimate population-level testing and case rates by race and ethnicity and may bias comparisons across subpopulations when information is not missing completely at random [18].

Missed infections occur when persons infected with SARS-CoV-2 are not documented in testing databases. Current evidence suggests that 60% of cases in the US were unreported during the first year of the pandemic [19]. The reasons for this are myriad. A large proportion of SARS-CoV-2 infections are asymptomatic [20]. Access to laboratory testing throughout the pandemic has been variable and often limited [21,22]. Furthermore, racial and ethnic disparities in COVID-19 testing stemming from structural racism have been documented [11]. For example, a study demonstrated the extent to which Black and Hispanic residents living in highly segregated US cities had lower access to COVID-19 testing sites in the early months of the pandemic [23]. Inequities in testing will be propagated into testing databases, resulting in selection bias and hindering our ability to quantify the true toll of the pandemic among certain populations.

In this study, we investigate racial and ethnic disparities in SARS-CoV-2 testing and cases in Holyoke, Massachusetts from March 8 to December 31, 2020. We address potential bias due to missing race and ethnicity data through multiple imputation and investigate missed infections and the impact of selection bias by comparing routinely collected testing data with a population-representative seroprevalence study of antibodies to COVID-19 conducted over the study period in Holyoke [24].

## METHODS

### Study setting

Holyoke is a post-industrial city in western Massachusetts with a population of 40,241 in 2019 [25]. Holyoke is in Hampden County, which has the highest level of social vulnerability in Massachusetts per the US Centers for Disease Control and Prevention (CDC)’s social vulnerability index (SVI), an index that uses US census data to determine the relative potential negative effects on census tracts caused by external stress on human health such as disease outbreaks [26]. Over half of the population identifies as Hispanic or Latino/a/x, while the remaining population is predominantly non-Hispanic white (Table 1). The non-Hispanic white community primarily resides in areas of lower SVI farther from the city center, with Hispanic or Latino/a/x populations residing in the city center in areas of higher SVI (Figure 1, Supplemental Table 1) [27]. We use the term “Hispanic” to refer to the “Hispanic or Latino/a/x” population for the remainder of the paper.

**Table 1.** Holyoke demographic distribution compared to original and imputed testing datasets (March 8, 2020 – December 31, 2020)

|  | Holyoke (ACS,<br>2019)<br>*n* (%) | Citywide<br>Testing:<br>Original<br>*n* (%) | Citywide Testing:<br>Imputed<sup>2</sup><br>*n* (%) | P-value<sup>6</sup> |
| --- | --- | --- | --- | --- |
| **Overall** | 40241 | 19658 | 19658 |  |
| **Race and ethnicity**<br>**category** |  |  |  |  |
| Hispanic | 21704 (53.9) | 8592 (56.8) | 10408 (52.9) | 0.026 |
| Non-Hispanic |  |  |  |  |
| White | 16636 (41.3) | 5880 (38.9) | 7917 (40.3) | 0.015 |
| Black | 1162 (2.9) | 436 (2.9) | 737 (3.8) | <0.001 |
| Asian | 239 (0.6) | 110 (0.7) | 299<sup>1</sup> (1.5) | <0.001 |
| Grouped race<sup>3</sup> | 500 (1.2) | 109 (0.7) | 296 (1.5) | 0.014 |
| *Missing* | -- | 4531 (23.0%) | -- | -- |
| **Age** |  |  |  |  |
| 0-19 | 10382 (25.8) | 3754 (19.1) | 3757 (19.1) | <0.001 |
| 20-44 | 14366 (35.7) | 8001 (40.7) | 8007 (40.7) | <0.001 |
| 45-59 | 7606 (18.9) | 3996 (20.3) | 3999 (20.3) | <0.001 |
| 60-84 | 7002 (17.4) | 3555 (18.1) | 3557 (18.1) | 0.012 |
| 85+ | 885 (2.2) | 338 (1.7) | 338 (1.7) | <0.001 |
| *Missing* | -- | 14 (<1%) | -- | -- |
| **Gender<sup>4</sup>** |  |  |  |  |
| Female | 20764 (51.6) | 10871 (55.3) | 10871 (55.3) | <0.001 |
| Male | 19477 (48.4) | 8732 (44.4) | 8732 (44.4) | <0.001 |
| Grouped gender<sup>5</sup> | -- | 55 (<1%) | 55 (<1%) | -- |
| **Census Tract Category** |  |  |  |  |
| High Hispanic | 10701 (26.6) | 5333 (27.1) | 5333 (27.1) | 0.001 |
| Medium Hispanic | 20413 (50.7) | 9949 (50.6) | 9949 (50.6) | 0.006 |
| Low Hispanic | 9127 (22.7) | 4376 (22.3) | 4376 (22.3) | 0.845 |
| **Social Vulnerability Index** |  |  |  |  |
| < 75<sup>th</sup> percentile in<br>Massachusetts | 9127 (22.7) | 4376 (22.3) | 4376 (22.3) | 0.845 |
| ≥ 75<sup>th</sup> percentile in<br>Massachusetts | 31114 (77.3) | 15282 (77.7) | 15282 (77.7) | 0.845 |

<sup>1</sup> Number of individuals classified as non-Hispanic Asian is slightly higher than the Holyoke population (based on ACS 2019).
<sup>2</sup> *n* (%) are averaged across M=20 imputed datasets. No measure of uncertainty shown for brevity.
<sup>3</sup> Grouped categories group includes other, two or more races, and American Indian or Alaskan Native
<sup>4</sup> The information listed for the Holyoke is the “Sex” variable from the ACS 2019 as that was the only question available.
<sup>5</sup> “Grouped gender” includes persons identifying as Transgender and persons with Unknown gender. No formal comparison available as ACS 2019 only includes “Male” and “Female”.
<sup>6</sup> P-values correspond two-sided one-sample t-tests with the null hypothesis that the observed proportion in the imputed testing data is equal to the Holyoke population proportion, and the variance estimated using Rubin’s rules. A statistically significant p-value for a particular group suggests that this group is under or over represented in the testing data relative to the Holyoke population.

**Figure 1.**
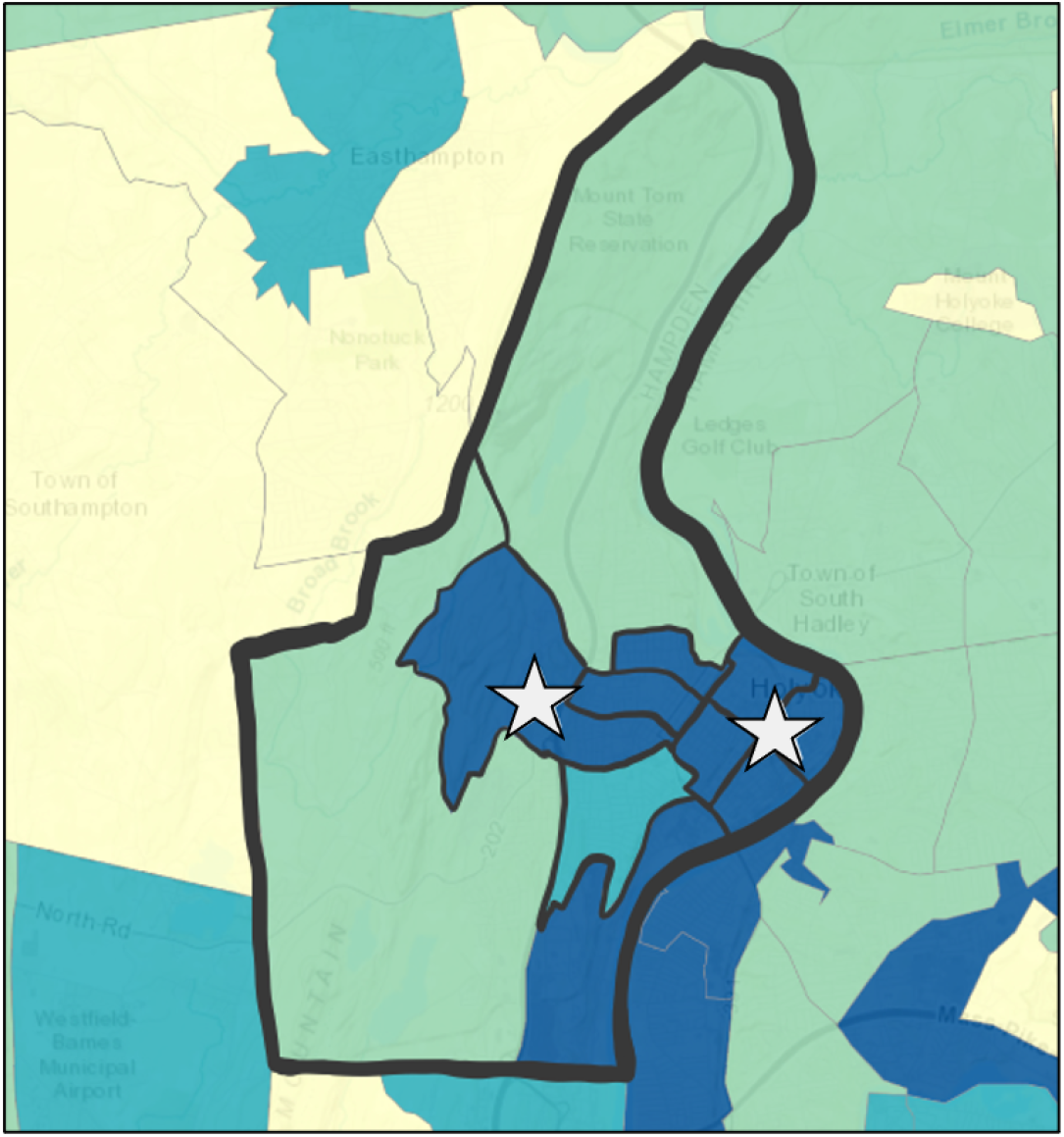
Holyoke census tracts shaded by social vulnerability index with dark blue indicating “high” vulnerability (>75^th^ percentile), light blue indicating “moderate to high” (50-75^th^), light green indicating “moderate to low” (25-50^th^), and yellow indicating “low” (<25^th^). Freely available “stop the spread” testing sites are shown by the stars. Figure was extracted and modified from CDC’s SVI Interactive Map at https://svi.cdc.gov/map.html.

### Data sources

The main data source in this study is the Holyoke COVID-19 testing database, which includes individual-level information on all COVID-19 tests of individuals residing in Holyoke, Massachusetts. The city-level COVID-19 testing data was managed by Massachusetts Virtual Epidemiologic Network (MAVEN), the state’s epidemiologic data system [28]. The data extracted for this analysis did not contain names or phone numbers.

We also utilized data from a seroprevalence survey of COVID-19 conducted from November 6 to December 31, 2020 among residents from a simple random sample of Holyoke households [24]. The study collected demographic information in addition to blood samples for serologic testing of COVID-19 antibodies from 328 individuals. The seroprevalence survey only reported seroprevalence estimates among Hispanic and non-Hispanic white groups due to small sample sizes among other racial and ethnic groups. Lastly, we used an address list for Holyoke (current as of June 2020) to map addresses to census tracts. This list was also used for household sampling in the seroprevalence study.

This study received approval from the Mass General Brigham Institutional Review Board (Protocol #2021P001714) and the Harvard Institutional Review Board (IRB20-1300).

### Study population

We utilized all available data from March 8, 2020 through March 8, 2021 (26,487 individuals with 97,049 tests) to impute missing data. After imputation, to facilitate comparisons between Holyoke testing data and the seroprevalence survey, we excluded testing data for Holyoke residents who did not receive at least one reverse transcriptase polymerase chain reaction (PCR) test between March 8 and December 31, 2020. We removed individuals residing in the ten long-term care facilities that were also excluded from the seroprevalence survey. Our final sample size included 19,658 individuals with 50,075 tests. See Supplemental Materials for details regarding data cleaning.

### Key Variables

Non-missing data from the testing database were leveraged to impute missing race and ethnicity. We derived variables related to the individual, the individual’s residence, and the individual’s test provider(s) (i.e. the name of the individual(s) who ordered the COVID-19 test).

Individual-level variables constructed from the testing data included: 1) Age at first test, categorized as <19, 20-44, 45-59, 60-85, and 85+ years. 2) Gender, categorized as male, female, and grouped gender. Grouped gender included individuals identifying as transgender or with unknown gender, and was created because of insufficient information to appropriately impute unknown gender and because the small number of self-identified transgender individuals (<10) precluded valid inference. 3) Race, collapsed into white, Black, Asian, and grouped race (other, two or more races, Native Alaskan or American Indian, Native Hawaiian or Pacific Islander) and Hispanic ethnicity. Grouped race was created for categories where the small sample size precluded valid inference or imputation convergence. A combined race and ethnicity categorical variable was constructed for analyses: Hispanic (any race), non-Hispanic white, non-Hispanic Black, non-Hispanic Asian, and non-Hispanic grouped race (Supplemental Materials).

Individual-level variables also included individuals’ testing characteristics. We created a variable for total number of PCR tests received and the number of weeks between March 8, 2020 and the individuals’ first COVID-19-related test, including PCR, rapid antigen, or antibody tests. We also created indicator variables for whether the individual ever had a positive PCR test result, received an antibody test and received an antigen test.

We analyzed four residence variables. First, an indicator for living in an apartment was created based on the existence of apartment numbers in the street address. Second, we mapped addresses to their respective census tracts and constructed a three-level categorical variable based on the proportion of individuals in each census tract identifying as “Hispanic” in the 2019 census: 1) high Hispanic proportion (>75%), 2) medium Hispanic proportion (25-75%), and 3) low Hispanic proportion (<25%). Third, using the census-address mapping, we constructed a binary variable indicating high social vulnerability (cutoff of 75^th^ percentile) based on the CDC’s SVI. Finally, we created a household identifier that grouped individuals with the same addresses.

Finally, we created variables to capture the distribution of race and ethnicity for a given testing provider, known as the *testing provider race and Hispanic ethnicity majority variables*, as testing providers who order tests for a large proportion of patients with a given race or ethnicity (determined through comparison to the Holyoke census population distribution) could be informative for imputing missing race and ethnicity data. See the Supplemental Materials for details on the construction of these variables.

### Outcome measures

Outcome measures were the positivity rate, case rate, and number of missed infections. *Positivity rate* is the number of people with at least one positive PCR test divided by the number of people who received at least one PCR test. *Case rate* refers to the number of people with at least one positive PCR test divided by the population of Holyoke. The number of *missed infections* is the difference between the case rate and the true SARS-CoV-2 infection rate, which is estimated from the seroepidemiologic survey. All outcomes correspond to March 8 through December 31, 2020.

### Statistical methods

#### Multiple imputation to address missing race and ethnicity data

For individuals with missing race and/or ethnicity in the testing data, we conducted multi-level multiple imputation to account for clustering by household of residence using the R jomo package [29]. jomo uses a joint modelling approach to impute missing information, and incorporates clustering through random intercepts. Addressing the clustering of race and ethnicity enables us to utilize the racial and ethnic make-up of an individual’s household when imputing their missing information. The completely observed covariates included in the imputation model were the number of PCR tests; indicators for prior positive PCR test result, prior antibody test, and prior antigen test; time to first test (weeks); household type (house, apartment, long-term care facility); census tract category; gender; and testing provider race and Hispanic ethnicity majority variables. Race and ethnicity were imputed separately, then combined into the race and ethnicity categorical variable described previously. The small number missing age values were also imputed.

We created twenty imputed datasets and used Rubin’s rules to obtain variance estimates for all estimates obtained in the multiple imputation procedure. As the testing data is a population-level data source, the only uncertainty comes from the imputation procedure itself, such that the within-imputation variance term in Rubin’s rules vanishes.

#### Assessing disparities in testing, accounting for missing race and ethnicity data

We compared the distribution of race and ethnicity, age, gender, and SVI between the Holyoke population (from the 2019 American Community Survey (ACS) [27]) and the individuals who received at least one COVID-19 test in the imputed testing data, to assess disparities in testing by demographic characteristics. This was done for each variable/category by conducting two-sided one-sample t-tests with the null hypothesis that the observed proportion in the imputed testing data is equal to the Holyoke population proportion, and the variance estimated using Rubin’s rules. Next, we investigated the impact of missing data in the testing database by comparing the distribution of these characteristics between the original and imputed testing data. Finally, using the imputed data, we compared testing characteristics (number of PCR tests, days to first PCR test, received antibody test) by race and ethnicity categories for individuals in the testing dataset.

#### Assessing disparities in infection, accounting for missing race and ethnicity data

We computed the positivity and case rate for each race and ethnicity category among the original testing population (excluding or separating individuals with unknown race and ethnicity category) and the imputed testing population (all individuals who received a test in Holyoke). This enables a comparison in rates between what is typically shown on COVID-19 data dashboards (“original”), which separates (for positivity rates) or excludes (for case rates) individuals with missing race and/or ethnicity, and what would be expected if there was no missing data (“imputed”). To examine the impact of missing data on disparities in infection, we also computed the positivity and case rate ratios between the Hispanic and non-Hispanic white population using both the imputed and the original testing data. A rate ratio over 1 suggests that the positivity (case) rate in the Hispanic population is higher than that in the non-Hispanic white population. To asses the impact of missing data by where individuals live, we repeated the above analysis by SVI.

#### Assessing disparities in missed infections, accounting for missing race and ethnicity data and potentially disparate testing rates

We computed the number of missed infections per 1000 individuals in each race and ethnicity category by taking the difference between the case rate from the imputed testing data and the estimated seroprevalence from the seroepidemiologic study. Seroprevalence is the proportion of the population with IgG antibodies and represents the best estimation for the proportion of the Holyoke population with a SARS-CoV-2 infection by December 31, 2020. Missed infections may be preferable over other metrics to assess disparities because this quantity incorporates disparities in case rates and disparities in testing. That is, missed infections may occur in the presence of disparate case rates and equal testing rates, equal case rates and disparate testing rates, or both. If case rates are disparate, testing rates must be correspondingly higher in the most-affected group to avoid missed infections. We constructed 95% credible intervals (CI’s) for the rate of missed infections in each race and ethnicity category using a parametric bootstrap procedure. Again, we repeated the above analysis by SVI to determine the impact of missed infections by location of residence.

## RESULTS

In the Holyoke testing population, 23.0% individuals had an unknown race and ethnicity category (n=4531), with 21.8% (n=4286) missing race, 19.7% (n=3869) missing ethnicity, and <1% missing age (n=14) (Table 1). Individuals with missing race and/or ethnicity were more likely to reside in low SVI census tracts, be under the age of 45 or over 84, have a lower total number of PCR tests, have an earlier week of first test, and live in a house vs an apartment. These individuals were less likely to have had an antibody test or to have ever tested positive (Supplemental Table 2).

After imputing race and ethnicity, the size of the non-Hispanic Asian population in the imputed testing data was slightly larger than the Holyoke population, leading to a significantly larger proportion of non-Hispanic Asian individuals in the imputed testing data. Further, we found significant, but not meaningful differences (all <1%) in all racial and ethnic categories between individuals who received at least one test and the Holyoke population. The distribution of SVI was similar between the Holyoke population and the imputed testing data. Individuals under 19 years and males were underrepresented in the imputed testing data compared to the city population, while individuals aged 20-44 were overrepresented.

### Disparities in testing

Testing rates were similar between non-Hispanic white and Hispanic populations with 476 compared to 480 per 1,000 individuals receiving at least one test by December 31, 2020. The median number of tests per individual was two with 49% of the Holyoke population having received at least one test during this period; there was no difference between the Hispanic and the non-Hispanic white population in terms of number of tests. Among those who received a test, the median time to first PCR test was 213 days from March 8, 2020, with Hispanic individuals having slightly longer time to first test (222 vs. 204 days among non-Hispanic white individuals) (Table 2). Antibody tests were most common among the non-Hispanic white population (3.2% had received an antibody test), while only 1.5% of Hispanic individuals received an antibody test (Table 2). There were no discernible trends by census tracts or density of Hispanic population in the number of PCR tests received over time (Figure 2).

**Table 2.** Holyoke testing characteristics by race/ethnicity category from citywide testing imputed dataset (March 8, 2020 – December 31, 2020)

|  | Overall | Hispanic | Non-Hispanic |  |  |  |
| --- | --- | --- | --- | --- | --- | --- |
|  |  |  | White | Black | Asian | Other |
| **Number of PCR tests per individual**<br>*median (IQR)*<sup>1</sup> | 2<br>(1, 3) | 2<br>(1, 3) | 2<br>(1, 3) | 1<br>(1, 2.46) | 1<br>(1, 2) | 1<br>(1, 2.64) |
| **Days to first PCR test**<br>*median (IQR)*<sup>1</sup> | 213<br>(145, 256) | 222<br>(151, 258) | 204<br>(143, 252) | 197<br>(128, 245) | 199<br>(125, 244) | 197<br>(130, 246) |
| **Ever received antibody test**<br>*n (%)*<sup>1</sup> | 428<br>(2.2) | 155<br>(1.5) | 250<br>(3.2) | 10<br>(1.4) | 8<br>(2.7) | 5<br>(1.7) |

<sup>1</sup> Statistics are taken within an imputed dataset and then averaged across the M=20 imputed datasets. No measures of uncertainty shown for brevity.

**Figure 2.**
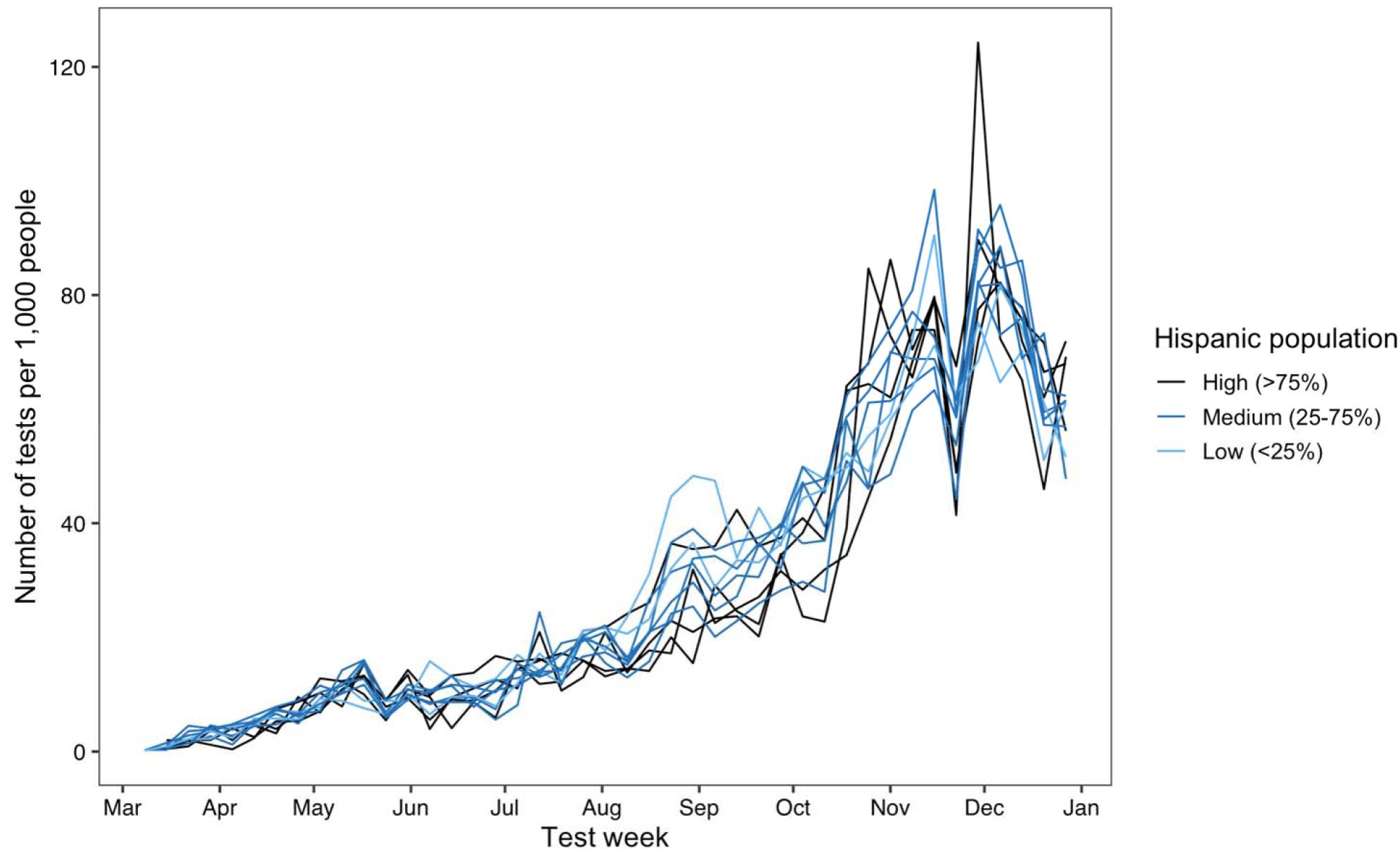
Weekly number of PCR tests per 1,000 population with each census tract. Each line represents a census tract and color represents low, medium, or high Hispanic population based on ACS 2019.

### Disparities in infection

The SARS-CoV-2 test positivity rate between March 8 and December 31, 2020 was 12.6%, corresponding to a case rate for Holyoke of 6.2%. The Hispanic population had higher positivity (16.9%) and case (8.1%) rates compared to the non-Hispanic white population (7.8% and 3.7%, respectively). Analyses using testing data with a separate category for “unknown” race or ethnicity, yielded a higher positivity rate across all race and ethnicity categories compared to the imputed population (Table 3). Notably, the positivity rate among the unknown group was lower than the complete case Holyoke population (3.4% vs. 15.4%). The case rates, while lower in the original testing data compared to those in the imputed population, were similar, as individuals with at least one positive SARS-CoV-2 test were less likely to have missing race and ethnicity than tested individuals who never had a positive SARS-CoV-2 test (7% and 12% missing, respectively); this is likely due to follow-up contact tracing efforts among confirmed cases. When computed using the imputed data, both the positivity and case rate ratios between Hispanic and non-Hispanic white individuals were 2.2 (2.0, 2.4), which was close to the estimated computed using original testing data (Table 3). Findings stratified by SVI were similar (Supplemental Table 3).

**Table 3.** Rates of SARS-CoV-2 infection from citywide testing original and imputed data (March 8, 2020 – December 31, 2020)

|  | Positivity rate |  | Rate ratio<sup>4</sup> |  | Case rate<sup>1</sup> |  | Rate ratio<sup>4</sup> |  |
| --- | --- | --- | --- | --- | --- | --- | --- | --- |
|  | Original | Imputed<sup>3</sup> | Original | Imputed<sup>5</sup> | Original | Imputed<sup>3</sup> | Original | Imputed<sup>5</sup> |
| **Overall** | **15.4** | **12.6** |  |  | **5.8** | **6.2** |  |  |
| Hispanic | 19.6 | 16.9<br>(16.7,17.0) | 2.1 | 2.2<br>(2.0, 2.4) | 7.8 | 8.1<br>(8.0, 8.1) | 2.3 | 2.2<br>(2.0, 2.4) |
| Non-Hispanic |  |  |  |  |  |  |  |  |
| White | 9.5 | 7.8<br>(7.7,7.9) | -- | -- | 3.4 | 3.7<br>(3.6, 3.8) | -- | -- |
| Black | 10.8 | 7.7<br>(7.0,8.3) |  |  | 4.0 | 4.9<br>(4.5, 5.2) |  |  |
| Asian<sup>2</sup> | 13.6 | 6.7<br>(5.1,8.4) |  |  | 6.3 | 8.4<br>(6.6, 10.2)<sup>2</sup> |  |  |
| Grouped race | 22.9 | 10.9<br>(8.7,13.1) |  |  | 5.0 | 6.4<br>(5.2, 7.7) |  |  |
| Unknown | 3.4 | -- |  |  | -- | -- |  |  |

<sup>1</sup> Number of cases per 100 persons <sup>2</sup> Number of individuals classified as non-Hispanic Asian is slightly higher than the Holyoke population (based on ACS 2019), potentially leading to inflated case rates <sup>3</sup> 95% confidence intervals constructed using Rubin's rules <sup>4</sup> Rate ratio compares Hispanic to non-Hispanic white population <sup>5</sup> 95% credible intervals obtained from a parametric bootstrap procedure

### Disparities in missed infections

To understand how any disparities in testing could have influenced case detection, we compared the case rates derived from the testing data to the seroprevalence survey. The prevalence of SARS-CoV-2 infection as measured by IgG antibodies was estimated to be 13.1% (6.9%, 22.3%) in the seroprevalence survey compared to 6.2% case rate in the testing data, meaning that an estimated 51.8% (9.2%, 71.9%) of Holyoke SARS-CoV-2 infections were not captured in the testing data [24]. This represents about 2,682 missed SARS-CoV-2 infections, or 67 missed infections per 1,000 people (Table 4). Among the Hispanic population, the estimated seroprevalence was 16.1% (6.2%, 31.8%) compared to an 8.1% case rate in the testing data, representing about 77 (0, 223) missed SARS-CoV-2 infections per 1,000 people. Among the non-Hispanic white population, the estimated seroprevalence was 9.4% (4.6%, 16.4%) compared to 3.7% in the testing data, representing about 58 (8, 124) missed infections per 1,000 people. There was a higher rate of missed cases overall, as well as a larger difference in Hispanic and non-Hispanic white-specific rates, among residents of high vs low SVI census tracts (Supplemental Table 4).

**Table 4.** Holyoke missed SARS-CoV-2 infections comparing case rates from imputed citywide testing and seroprevalence from seroepidemiologic study data (March 8, 2020 – December 31, 2020)

|  | Citywide Testing<br>Imputed<sup>1</sup> | Sero-<br>epidemiologic<br>study<sup>2,3</sup> | Number of<br>missed cases<br>per 1,000<br>people<sup>2,3,4</sup> |
| --- | --- | --- | --- |
| Overall | 6.2 | 13.1<br>(6.9, 22.3) | 68.5<br>(6.5, 171.1) |
| Hispanic | 8.1<br>(8.0, 8.1) | 16.1<br>(6.2, 31.8) | 79.4<br>(0, 239.7) |
| Non-Hispanic |  |  |  |
| White | 3.7<br>(3.6, 3.8) | 9.4<br>(4.6, 16.4) | 59.9<br>(8.6, 125.7) |
| Black | 4.9<br>(4.5, 5.2) | -- | -- |
| Asian | 8.4<br>(6.6, 10.2) | -- | -- |
| Grouped race | 6.4<br>(5.2, 7.7) | -- | -- |

<sup>1</sup> Uncertainty intervals are (2.5<sup>th</sup>, 97.5<sup>th</sup>) percentiles across 20 imputed datasets
<sup>2</sup> Seroprevalence estimates only available for Hispanic and non-Hispanic White groups
<sup>3</sup> Uncertainty intervals are 95% credible intervals
<sup>4</sup> Lower credible interval range truncated at 0 as negative number of missed infections is not possible

## DISCUSSION

In this study, we combined routinely collected public health data with rigorous statistical methodology and a representative seroprevalence survey to identify disparities in SARS-CoV-2 testing and case rates by race and ethnicity in Holyoke, MA. We highlighted how missing data (due to an absence of race/ethnicity data or undetected infection) may bias these estimates.

The positivity and case rate among Holyoke’s Hispanic population were nearly double that of the non-Hispanic white population, revealing a disproportionate burden of SARS-CoV-2 in this population. Other studies have similarly found higher risk of infection among the US Hispanic population [5-8]. Testing rates were similar by race and ethnicity. This finding contrasts with other studies in Massachusetts and nationally [21-23]. Similar testing rates may be explained by the presence of two “Stop the Spread” sites, public, free-of-charge mass-testing sites deployed in Massachusetts during the early phases of the pandemic. However, equality in testing rates does not necessarily translate to equitable testing. Higher positivity and case rates indicate a need for a higher testing rate in the Hispanic population. A higher rate of missed infections in this population, suggests that, though equal, the rate of testing in the Hispanic population was inadequate given the greater burden of infection. Missed SARS-CoV-2 infections preclude opportunities to reduce onward transmission, and contribute to the spread of infections within these communities, further driving inequities. If perpetuated into the era of ‘test and treat’, missed infections may result in missed opportunities to reduce morbidity and other mortality. The reasons underlying this testing inequity likely stem from structural barriers, that limit access to healthcare even when health sites are present such as: incomplete language accessibility of messaging related to Covid-19 testing, including messaging regarding cost and insurance requirements, inflexible employer sick leave policies contributing to fear of testing positive, fear of deportation amongst undocumented individuals, and long wait times at testing centers, even at Stop the Spread sites where demand was heightened by the influx of people coming from neighboring locales to get tested [30].

Positivity rates were higher and case rates were lower in the complete cases testing data compared to those in the imputed data, due to the lower rate of missing race and ethnicity information among cases compared to tested individuals who never had a positive SARS-CoV-2 test. Although our estimates of *disparities* in infection rates (positivity/case rate ratios) were not meaningfully impacted by missing data, this is due to the nature of missingness in this study; other studies have shown a substantial impact on findings, underscoring the importance of appropriately accounting for missing data to more accurately inform targeted public health responses [18, 31].

Over half of the cases were not captured by the testing data, highlighting the role that bias may play if public health officials focus on testing data only [32-33]. Case rates from testing data alone will greatly underestimate the true burden of SARS-CoV-2 infections. Testing data may still be useful for comparing health outcomes between groups if the testing population is representative of the total population. If not representative, the presence of selection bias should be evaluated and addressed [32-35].

We utilized a multi-level multiple imputation procedure that allowed us to impute race and ethnicity separately, and to account for clustering of race and ethnicity by household. Two potential limitations of this procedure are that it assumes that the data are missing at random (MAR) [29], and that the imputation model is correctly specified; when these assumptions do not hold, resulting estimates may be biased. All variables included our imputation model were significantly associated with missingness in the race and ethnicity variable (Supplemental Table 2), though we did not have access to other potentially informative data such as individuals’ occupation, which may threaten the MAR assumption. Future health data systems would benefit from collecting more information on characteristics known to be associated with race and ethnicity. Other imputation procedures such as Bayesian Improved Surname Geocoding (BISG) [18,36] or population calibrated multiple imputation (PCMI) [37] could be employed, however, these require access to individuals’ surnames or knowledge of the true testing race and ethnicity distribution, respectively, neither of which we had access to in the current study. Ultimately, the most appropriate strategy will depend on the available information.

This study has several limitations: First, we grouped American Indian and Alaskan Native and Native Hawaiian and Pacific Islander into a “grouped” race category because the number of individuals was very small. Grouping race categories this way will mask true differences between groups and may further marginalize these communities [38]. Second, there was no standardized method of collecting race or ethnicity information at testing facilities. Prior research has shown that differences in the ordering and question format for race and ethnicity data can result in inconsistent answers [39, 40]. Misclassifications at the outset would propagate into the multiple imputation procedure. We attempted to mitigate this through our data cleaning process, which was informed by discussions with Holyoke testing sites about how race and ethnicity data was collected. Third, there was no standardized method of collecting gender at testing facilities. The testing data only had options for “male”, “female”, and “transgender” with only 6 (0.03%) individuals listed as transgender, over ten-fold smaller than the estimated percentage of transgender individuals living in Massachusetts [41]. This could be due to either a disparity in testing by gender or an artefact of non-standardized data collection on gender. These data issues can be obviated by instituting standardized data collection procedures at testing facilities. Finally, the seroprevalence survey used to estimate the true infection rates was itself subject to several limitations [24]; in this study, these manifested as insufficient data to estimate missed infection rates among all race and ethnicity groups, and wide CIs in groups where estimation was possible. Despite these limitations, our analysis can serve as an example of how to investigate where and how racial and ethnic disparities may lead to differential testing rates, case rates, and missed infections.

Routinely collected testing data is a vital resource for targeting public health responses. In this study, we highlight a disproportionate burden of SARS-COV-2 infections among the Hispanic population in Holyoke and an inequity in testing between Hispanic and non-Hispanic white populations. We address biases inherent to analyses using routinely collected testing data by using multiple imputation of missing data and comparing the testing data to a representative seroprevalence survey. While the statistical procedures presented in this paper enhance rigor, they are no substitute for consistent data quality and coordinated, integrated data systems, which are needed to uncover the true burden of the COVID-19 pandemic by demographic characteristics, and to guide an equitable response to the pandemic.

## Data Availability

All data produced in the present study are available upon reasonable request to the authors

## Acknowledgements

We thank the rest of the Holyoke Board of Health, who supported this study. We are also grateful to Scott Troppy and Reed Sherrill from the Massachusetts Department of Public Health for their assistance with organizing and understanding the primary data source.

