## Supplemental Materials for "Missing data and missed infections: Investigating racial and ethnic disparities in SARS-CoV-2 testing and infection rates in Holyoke, Massachusetts"

*Supplemental Material*

**Address cleaning and census tract matching**

109,964 tests^*^ corresponding to 34,106 individuals

^*^From March 8, 2020 to March 8, 2021

Extract city, state, and zip code from full addresses.

Remove 7,643 tests (1,99 individuals) corresponding to addresses outside of Holyoke, MA.

103,235 tests corresponding to 32,115 individuals

98,609 tests corresponding to 30,485 individuals

Remove 3,726 tests (1,630 individuals) corresponding to missing or unknown addresses.

Separate out house number and street name from full addresses, carry out manual updates to street names:

- change street, drive, highway, avenue etc. into common abbreviations (st, dr, hwy, ave)
- correct spelling errors

Separate out apartment numbers from street name indicating floor, unit, apartment, suite (including common misspellings and abbreviations)

Match cleaned street/house number combinations to Holyoke address census

- among unmatched combinations, use the stringdist R package for approximate string matching with distance of five
- among remaining unmatched combinations, use censusxy R package to identify census tract

Remove 798 (346 individuals) corresponding to addresses that could not be matched

Overall, removed:

- 7,643 (6.9%) of tests with address outside of Holyoke
- 3,726 (3.4%) of tests with missing addresses
- 798 (0.7%) of tests with unreconciled addresses

Results in 97,797 tests corresponding to 30,132 individuals

**De-duplication (to create individual-level dataset):** Each test has a unique case ID that can be used to link multiple tests to one individual. When a new laboratory test is entered into the MAVEN system, it is matched against all existing individuals’ full name, date of birth, and address. If any of the variables are off by a single letter or number, a new case ID will be generated. As this process is quite stringent and likely misclassifies tests to new individuals, we performed an additional de-duplication step with our cleaned addresses:

- Match individuals based on cleaned address, gender, and date of birth.
- If the matching yields a grouping of IDs associated with different race or ethnicity values, separate these out with the following exceptions:
  - The number of unique race values is two, one of which is “unknown”, and there is only 1 Hispanic value.
    - Assign all “unknown” values to the other specified value.
  - The number of unique race values is two, one of which is ‘unknown’ and there are two Hispanic values, but one of them is ‘unknown’.
    - Assign all “unknown” values to the other specified value.
  - The number of unique race values is three, but two of these are “other” and “unknown”, and Hispanic is either 1) one unique value or 2) two unique values, with one of them equal to “unknown”.
    - Assign all “unknown” and “other” values to the other specified value.

This results in 97,797 tests corresponding to 26,851 individuals (compared to 30,132 individuals prior to de-duplication*). At this point, we remove tests corresponding to individuals that never received a PCR test (i.e. they only have antibody OR antigen tests in the testing data). This results in 97,049 tests corresponding to 26,487 individuals.*

**Cleaning demographic variables:** Collection of race, ethnicity, and gender were not standardized across testing sites, such that the question wording, ordering of questions, and collection method (e.g. verbal or written) may have varied over time and across testing sites. Further, if an individual had more than one test or encounter with the MAVEN data reporting (e.g. contact tracing), additional race(s) could be appended to their unique case ID, sometimes resulting in conflicting information. We performed the follow steps to clean demographic variables before imputation and analysis:

**Race:** (multiple race categorizations appear separated by commas, which may be due to user input, but also due to repeat testing or contact tracing efforts)

- If only one race is listed, that race is kept
- If one race category is listed with “other” or “unknown”, then only the listed race is kept
- If more than one race listed, then categorized as “Two or more”
- If more than one race with “other” or “unknown”, then categorized as “Two or more”

**Hispanic or Latino/a/x Ethnicity:** (corresponds to two fields: “Hispanic, Spanish, or Latin origin?” and a drop-down menu of ethnic origin options)

- If “Hispanic, Spanish, or Latin origin?” is “Yes” then classified as “Hispanic or Latino/a/x”
- If “Hispanic, Spanish, or Latin origin?” is “No” or “Unknown” with no ethnic origin listed, then respectively classified as “Non-Hispanic or Latino/a/x” or “Unknown”
- If “Hispanic, Spanish, or Latin origin?” is “No” or “Unknown” and ethnic origin is not from a Hispanic, Spanish, or Latino origin country (as defined by the United States Census Bureau), then classified as “Non-Hispanic or Latino/a/x”
- If “Hispanic, Spanish, or Latin origin?” is “No” or “Unknown” and ethnic origin is from a Hispanic, Spanish, or Latino origin country (as defined by the United States Census Bureau), then classified as “Non-Hispanic or Latino/a/x”

**Accounting for small race and ethnicity categories before imputation:**

- If non-Hispanic or Latino/a/x ethnicity and “Other” race, then made “Unknown” as there were only three individuals in Holyoke (ACS 2019) with this classification
- If “Two or more”, “American Indian Alaskan Native” or “Native Hawaiian Pacific Islander”, then “Grouped” as these are all <1.0% of non-Hispanic Holyoke population

**Gender:** There was one “Gender” field with entries “Male”, “Female”, “Transgender”. We could not confirm that “Gender” was collected at every site (vs. “Sex”), and if individuals were always given the Male/Female/Transgender options, additional options for gender, or only the binary Male/Female. There were only six individuals identifying as “Transgender” in the testing dataset, precluding valid inference among this group of individuals. Thus, for imputation and analysis, we created a “Grouped” category combining “Transgender” and “Unknown”. *Future data collection on gender identity should be standardized across testing sites and allow individuals to self-describe.*

**Testing provider race and Hispanic ethnicity majority variables:** We created variables to capture the distribution of race and ethnicity for a given testing provider, known as the *testing provider race and Hispanic ethnicity majority variables*, as testing providers with a large proportion of patients with a given race or ethnicity (determined through comparison to the census population distribution) could be informative for imputing missing race and ethnicity data. For each provider in the dataset, we computed the observed proportion of individuals with at least one test ordered by the provider for each Hispanic, white, Black, Asian, and grouped race category. To determine if the testing provider had a large proportion of patients with a given race or Hispanic ethnicity, we conducted one sample tests of proportions with a one-sided alternative with the null values set to the Holyoke census population distribution of the Hispanic, white, Black, Asian, and grouped race categories (53.9%, 87.5%, 4.5%, 0.7%, 7.3% respectively). For each provider, we then constructed an indicator variable for Hispanic ethnicity and for each race category. These indicator variables were assigned a value of one if the null hypothesis for the one-sided test was rejected, the number of individuals with known race (or ethnicity) was at least 10, and less than 50% of the tested individuals had missing race (or ethnicity). We multiplied these indicator variables by the difference between the provider-specific observed proportion and the respective Holyoke census proportion. The provider variables for Hispanic ethnicity and each race category were then mapped back to, and summarized for, each individual; if an individual had multiple providers, the average of each of these variables was computed.

**Additional tables:**

**Table 1.** Distribution of race and ethnicity groups by census tract.

| **Census tract** | **SVI^1,2^** | **Hispanic**  (%) | **NH-white**  (%) | **NH-Black**  (%) | **NH-Asian**  (%) | **NH-Grouped^3^**  (%) |
| --- | --- | --- | --- | --- | --- | --- |
| **8114** (N=2335*)* | High | 2174 *(93.1)* | 80 *(3.4)* | 64 *(2.7)* | 0 *(0)* | 17 *(0.7)* |
| **8115**  (N=2197) | High | 1935 *(88.1)* | 95 *(4.3)* | 79 *(3.6)* | 0 *(0)* | 88 *(4.0)* |
| **8116**  (N=3952*)* | High | 3518 *(89.0)* | 314 *(7.9)* | 101 *(2.6)* | 0 *(0)* | 19 *(0.5)* |
| **8117**  (N=2217) | High | 1751 *(79.0)* | 404 *(18.2)* | 5 *(0.2)* | 36 *(1.6)* | 21*(0.9)* |
| **8118**  (N=4085) | High | 2616 *(64.0)* | 1367 *(33.5)* | 53 *(1.3)* | 10 *(0.2)* | 39 *(1.0)* |
| **8119**  (N=3603) | Low | 385 *(10.7)* | 3042 *(84.4)* | 131 *(3.6)* | 42 *(1.2)* | 3 *(0.1)* |
| **8120.01**  (N=3370) | High | 1938 *(57.5)* | 1201 *(35.6)* | 156 *(4.6)* | 44 *(1.3)* | 31 *(0.9)* |
| **8120.02**  (N=4590) | High | 2289 *(49.9)* | 1999 *(43.6)* | 178 *(3.9)* | 35 *(0.8)* | 89 *(1.9)* |
| **8121.01**  (N=5524) | Low | 883 *(16.0)* | 4613 *(83.5)* | 28 *(0.5)* | 0 *(0)* | 0 *(0)* |
| **8121.03**  (N=3932) | High | 2293 *(58.3)* | 1337 *(34.0)* | 282 *(7.2)* | 0 *(0)* | 20 *(0.5)* |
| **8121.04**  (N=4436) | High | 1922 *(43.3)* | 2184 *(49.2)* | 85 *(1.9)* | 72 *(1.6)* | 173 *(3.9)* |

^1^U.S. Census Bureau (2019). American Community Survey, Demographic and Housing Estimates

^2^High SVI corresponds to SVI ≥75^th^ percentile in Massachusetts, low SVI corresponds to SVI <75^th^ percentile in Massachusetts

^3^Grouped categories group includes other, two or more races, and American Indian or Alaskan Native

**Table 2.** Characteristics associated with missingness in race and ethnicity categorical variable

| **Characteristic** | **Overall**  N=19658 | **Missing**  N=4531 (23.0%)^1^ | **P-value**^2^ |
| --- | --- | --- | --- |
| **Social vulnerability index** |  |  | 0.033 |
| ≥ 75^th^ percentile in Massachusetts | 15282 | 3470 (22.7) |  |
| < 75^th^ percentile in Massachusetts | 4376 | 1061 (24.2) |  |
| **Census tract category** |  |  | 0.086 |
| *Low Hispanic* | 4376 | 1061 (24.2) |  |
| *Medium Hispanic* | 9949 | 2244 (22.6) |  |
| *High Hispanic* | 5333 | 1226 (23.0) |  |
| **Age at first test** |  |  | <0.0001 |
| *0-19* | 3754 | 898 (23.9) |  |
| *20-44* | 8001 | 1955 (24.4) |  |
| *45-59* | 3996 | 840 (21.0) |  |
| *60-84* | 3555 | 746 (21.0) |  |
| *85+* | 338 | 86 (25.4) |  |
| *Missing* | 14 | 6 (42.9) |  |
| **Antibody test** |  |  | <0.0001 |
| *No* | 19266 | 4503 (23.4) |  |
| *Yes* | 392 | 28 (7.1) |  |
| **Antigen test** |  |  | 0.026 |
| *No* | 19363 | 4479 (23.1) |  |
| *Yes* | 295 | 52 (17.6) |  |
| **Ever positive** |  |  | <0.0001 |
| *No* | 17176 | 4377 (25.5) |  |
| *Yes* | 2482 | 154 (6.2) |  |
| **House type** |  |  | 0.015 |
| *Apartment* | 5266 | 1150 (21.8) |  |
| *House* | 14392 | 3381 (23.5) |  |
| **Gender category** |  |  | <0.0001 |
| *Female* | 10871 | 2397 (22.0) |  |
| *Male* | 8732 | 2105 (21.1) |  |
| *Unknown/transgender* | 55 | 29 (52.7) |  |
| **Number of PCR tests** |  |  |  |
| *1* | 8846 | 3374 (38.1) | <0.0001 |
| ≥ 1 | 10812 | 1157 (10.7) |  |
| **Week of first test** |  |  |  |
| *< 12* | 2136 | 640 (30.0) | <0.0001 |
| ≥ 12 | 17522 | 3891 (22.2) |  |

^1^In parentheses are the percentages of missing race and/or ethnicity for each characteristic shown

^2^P-values are obtained from Chi-squared tests of association for categorical variables

**Table 3.** Rates of SARS-CoV-2 infection from citywide testing original and imputed data (March 8, 2020 – December 31, 2020)

| **SVI** | **Race ethnicity** | **Positivity rate** | | **Rate ratio^3^** | | **Case rate** | | **Rate ratio^3^** | |
| --- | --- | --- | --- | --- | --- | --- | --- | --- | --- |
| **High** |  | Original | Imputed^2^ | Original | Imputed^4^ | Original | Imputed^2^ | Original | Imputed^4^ |
|  | Overall | 16.4 | 13.5 |  |  | 6.2 | 6.6 |  |  |
|  | Hispanic | 19.4 | 16.8  (16.6, 16.9) | 2.1 | 2.2  (2.0, 2.5) | 7.7 | 8.0  (7.9, 8.0) | 2.3 | 2.1  (1.7, 2.7) |
|  | NH-white | 9.3 | 7.5  (7.3, 7.7) | -- | -- | 3.4 | 3.8  (3.7, 3.9) | -- | -- |
| **Low** |  |  |  |  |  |  |  |  |  |
|  | Overall | 11.9 | 9.6 |  |  | 4.3 | 4.6 |  |  |
|  | Hispanic | 22.5 | 17.9  (17.3, 18.6) | 2.3 | 2.2  (1.9, 2.4) | 9.3 | 9.7  (9.4, 9.9) | 2.7 | 2.7  (2.2, 3.5) |
|  | NH-white | 9.7 | 8.3  (8.1, 8.4) | -- | -- | 3.4 | 3.6  (3.6, 3.7) | -- | -- |

**^1^** Number of cases per 100 persons

**^2^** 95% confidence intervals constructed using Rubin’s rules

**^3^** Rate ratio compares Hispanic to non-Hispanic white population

**^4^** 95% credible intervals obtained from a parametric bootstrap procedure

**Table 4.** Holyoke missed SARS-CoV-2 infections comparing case rates from imputed citywide testing and seroprevalence from seroepidemiologic study data (March 8, 2020 – December 31, 2020)

| **SVI** | **Race ethnicity** | Citywide Testing Imputed^1^ | Sero-epidemiologic study^2^ | Number of missed cases per 1,000 people^2,3^ |
| --- | --- | --- | --- | --- |
| **High SVI** | Overall | 6.6 | 14.4 (7.1,25.5) | 74.4  (3.5, 185.3) |
|  | Hispanic | 8.0 (7.9, 8.0) | 16.1 (6.2, 32.0) | 80.9  (0, 238.2) |
|  | Non-Hispanic white | 3.8 (3.7, 3.9) | 10.6 (4.3, 20.4) | 68.2  (3.1, 163.2) |
| **Low SVI** | Overall | 4.6 | 8.2 (3.1, 16.9) | 37.3  (0, 111.6) |
|  | Hispanic | 9.7 (9.4, 9.9) | 13.1 (2.8, 37.5) | 38.1  (0, 266.6) |
|  | Non-Hispanic white | 3.6 (3.6, 3.7) | 7.7 (2.7, 16.3) | 41.7  (0, 124.2) |

^1^Uncertainty intervals are (2.5^th^, 97.5^th^) percentiles across 20 imputed datasets

^2^ Uncertainty intervals are 95% credible intervals

^3^ Lower credible interval range truncated at 0 as negative number of missed infections is not possible
